# Pandemic modelling for regions implementing an elimination strategy

**DOI:** 10.1101/2022.07.18.22277695

**Authors:** Amy Hurford, Maria M. Martignoni, J.C. Loredo-Osti, Francis Anokye, Julien Arino, Bilal Saleh Husain, Brian Gaas, James Watmough

## Abstract

During the COVID-19 pandemic, some countries, such as Australia, China, Iceland, New Zealand, Thailand, and Vietnam successfully implemented an elimination strategy, enacting strict border control and periods of lockdowns to end community transmission. Atlantic Canada and Canada’s territories implemented similar policies, and reported long periods with no community cases. In Newfoundland and Labrador (NL), Nova Scotia, and Prince Edward Island a median of 80% or more of daily reported cases were travel-related from July 1, 2020 to May 31, 2021. With increasing vaccination coverage, it may be appropriate to exit an elimination strategy, but most existing epidemiological frameworks are applicable only to situations where most cases occur in the community, and are not appropriate for regions that have implemented an elimination strategy. To inform the pandemic response in regions that are implementing an elimination strategy, we extend importation modelling to consider post-arrival travel restrictions, and pharmaceutical and non-pharmaceutical interventions in the local community. We find that shortly after the Omicron variant had begun spreading in Canada, the expected daily number of spillovers, infections spread to NL community members from travelers and their close contacts, was higher than any time previously in the pandemic. By December 24, 2021, the expected number of spillovers was 44% higher than the previous high, which occurred in late July 2021 shortly after travel restrictions were first relaxed. We develop a method to assess the characteristics of potential future community outbreaks in regions that are implementing an elimination strategy. We apply this method to predict the effect of variant and vaccination coverage on the size of hypothetical community outbreaks in Mount Pearl, a suburb of the St. John’s metropolitan area in NL. Our methodology can be used to evaluate alternative plans to relax public health restrictions when vaccine coverage is high in regions that have implemented an elimination strategy. This manuscript was submitted as part of a theme issue on *“Modelling COVID-19 and Preparedness for Future Pandemics”*.

## Introduction

To manage SARS-CoV-2 infections, countries including Australia, China, Iceland, New Zealand, Thailand, and Vietnam used an *elimination approach* (also known as a zero-COVID policy). This approach combines strong border control to diminish travel-related cases with pharmaceutical (PIs) and non-pharmaceutical interventions (NPIs) that reduce or completely end community transmissions if border measures fail (Baker et al., 2020a; Heywood and Macintyre, 2020). Elimination differs from *eradication* in that its intended region of influence is localized, typically to the jurisdiction pursuing the goal. This policy was also used in infranational jurisdictions such as Atlantic Canada and Canada’s territories (Bignami, 2021; Contandriopoulos, 2021; Department of Health and Community Services, NL, 2022).

Until the end of 2021, countries that used an elimination strategy had less SARS-CoV-2 mortality (Baker et al., 2020b; Nam et al., 2020) and less stringent local restrictions when there were no community cases, which resulted in less psychological distress (Aknin et al., 2022). Regions that implemented elimination strategies may have also had stronger economies (König and Winkler, 2021). Newfoundland and Labrador (NL), which implemented a containment approach (Department of Health and Community Services, NL, 2022), achieved prolonged periods with no community cases and low SARS-CoV-2 mortality: 3.6 SARS-CoV-2 deaths per 100,000 people in NL, compared to 78.1 SARS-CoV-2 deaths per 100,000 people in Canada from the beginning of the pandemic until December 31, 2021 (NL: 19 deaths (Government of Newfoundland and Labrador, 2021a) for a provincial population of 521,854 people (Statistics Canada, 2021); Canada: 30,024 SARS CoV-2 deaths (Public Health Agency of Canada, 2022) for a national population of 38,426,473 (Statistics Canada, 2021). Success similar to that of NL occurred throughout Atlantic Canada and in Canada’s territories.

The feasibility of an elimination strategy depends on vaccine availability and uptake. Hong Kong had low numbers of SARS-CoV-2 cases through strict border control, quarantine and NPIs, but did not vaccinate abundantly, which exposed the population to severe disease outcomes when the Omicron variant emerged (Ma and Parry, 2022). The feasibility of an elimination strategy may depend on variant characteristics and jurisdictional geographic and social characteristics (Silver, 2022; Martignoni and Hurford, 2022; Department of Health and Community Services, NL, 2022). In early 2022, following the establishment of the Omicron variant, NL shifted from a containment to a mitigation approach (Department of Health and Community Services, NL, 2022), with many of the jurisdictions that had implemented an elimination strategy responding similarly (a notable exception is China who continued to pursue an elimination strategy). The elimination strategy is most likely appropriate *in specific locations and for specific periods of time* as the costs and benefits of the strategy likely depend on complex interactions between regional characteristics, public health policy, community behavioral responses, and variant epidemiological characteristics.

While implementing an elimination strategy, it is important to develop tools to assess the risk of community outbreaks, to evaluate whether border controls should be upscaled or released. In the following, we define an *importation* as an individual who arrives in the local jurisdiction from another jurisdiction while infected with SARS-CoV-2. An importation occurs when a traveler is infected at the point of origin, or during travel to their destination. A *travel-related* infection refers to both an importation and close contacts who become infected by the traveler. A *spillover* is an infection from an individual with a travel-related infection to a community member that is not a close contact of the traveler, and a *community* infection is when a community member is infected either as a spillover, or from another community member. We note that this terminology differs from that used in NL Public Service Advisories, which reported cases as ‘related to international (or domestic) travel’ or ‘close contacts of a known case’. Infections and cases differ in that cases are the infections that are reported.

Travel-related and community infections arise through different processes, and therefore carry different risks and occur at different rates. The rate of arriving importations is dependent on the prevalence of infection at the travelers’ points of origin, the risk of infection during travel, and the rates of inbound travel to the local community (Russell et al., 2021). The rate that travel-related infections generate subsequent infections depends on contact rates with community members and can be reduced through post-arrival travel restrictions (Arino et al., 2020; Chen et al., 2021; Dickens et al., 2020). When travelers are to self-isolate post-arrival, infections can be spread to household members. In regions with few community cases of SARS-CoV-2, it is necessary to distinguish between importations, close contacts who were infected by an imported infection, and community infections (see Price et al. 2020 for related comments).

Here, we develop an approach to estimate the potential future impact of SARS-CoV-2 in communities that have experienced long periods with a high percentage of cases that are travel-related. Many models focus on community spread, without distinguishing between travel-related and community cases, and are not suitable for this purpose. During the pandemic, new SARS-CoV-2 variants emerged (Otto et al., 2021), and our framework considers this evolving risk. Our approach uses two models in a pipeline, expected spillovers and community spread, rather than only a single model that couples both. Our first model predicts the expected number of community members that are infected by travelers (i.e., spillovers) and considers three categories of public health measures: post-arrival travel restrictions, NPIs in the local community, and vaccination. Our second model describes a hypothetical future community outbreak and considers different variants and levels of vaccine coverage. The first model, describing the expected number of spillovers, is not coupled to the second model, describing a community outbreak, because community outbreaks might hypothetically begin on any given day, and averages taken across hypothetical outbreak start dates obscure key information (Juul et al., 2021). Considering a pipelined uncoupled framework is useful because some decisions that public health officials make are conditional on whether a community outbreak has been detected (notably the implementation of NPIs as part of an elimination strategy), while other decisions are better informed by the average across community outbreaks with all possible hypothetical start dates (i.e., generally applicable measures, such as provincial mask mandates when surveillance is low and importations are frequent).

In July 2021, most Canadians had received at least one dose of a SARS-CoV-2 vaccine, and there was a need to transition to a sustainable approach for SARS-CoV-2 management should high immunity levels be maintained. After vaccination, continued isolation of regions that implemented elimination strategies might be unrealistic, particularly given the economic and social impacts of these strategies (Committee for the Coordination of Statistical Activities, 2021). At this time, there was a need to develop guidelines to advise regions with zero or low SARS-CoV-2 prevalence in exiting elimination strategies (Lokuge et al., 2021; Open Society Common Purpose Taskforce, 2021). This remains an important topic even as most Canadian provincial governments have relaxed COVID-19 control measures. Indeed, vaccine coverage still lags in a large proportion of the world, and the emergence risk of novel variants remains high (Otto et al., 2021).

## Materials and Methods

### Data

Our analysis combines data from multiple sources (summarized in Table 1) including the Public Health Agency of Canada (PHAC), and the Newfoundland and Labrador Centre for Health Information (NLCHI). A data source for travel-related cases was the COVID-19 Canada Open Data Working Group (CCODWG) (Berry et al., 2020, 2021), a group of volunteers who curated data from government and non-government sources. We validated the CCODWG data with travel-related cases as reported by the NL and NB provincial governments and found that the CCODWG data accurately describes the number of travel-related cases in NL and NB (Figure S1). Another data source was the Bank of Canada NPI stringency index (Cheung et al., 2021), which was used to measure the severity of NPIs implemented in NL.

**Table 1:** Data sources. The line list for CCODWG (Berry et al., 2020) was discontinued on May 31, 2021, and as such, no data on travel-related cases or close contacts of travelers are available from CCODWG after this date.

| Data source | Variables | Timeframe | Jursidictions | Figures |
| --- | --- | --- | --- | --- |
| COVID Canada Open Data Working Group | Travel-related cases (daily)<br>Close contact cases (daily) | July 1, 2020-May 31, 2021 | NB, PEI, NS, NL, YT, NWT | Figs. 1; 2; and S1 |
| Newfoundland and Labrador Centre for Health Information | Travel-related cases (daily) | July 1, 2020-December 24, 2021 | NL | Figs. 2; 3A,E; and S1 |
| Government of New Brunswick public releases | Travel-related cases (daily) | January 1, 2021-May 31, 2021 | NB | Fig. S1 |
| Public Health Agency of Canada public data | New cases (daily) | March 15, 2020-December 24, 2021 | All Canadian provinces | Explanatory variable for model fit in Fig. 2 |
| Public Health Agency of Canada public data | Variant frequency (weekly) | March 14, 2020-December 24, 2021 | Canada | Fig. 3A,E; and S2A |
|  | Vaccination levels (weekly) |  | Canada and NL | Fig. 3C,E; and S2B |
| Newfoundland and Labrador Centre for Health Information | Community cases (daily) | March 14, 2020-December 24, 2021 | NL | Fig. 3E |
|  | Close contacts infected per travel-related case |  |  | Fig. 3E |
|  | Number of cases and symptom onset date for the Mt. Pearl outbreak | February 8 – 24, 2021 |  | Fig. 4 |
| Bank of Canada | NPI stringency (daily) | July 1, 2020-December 24, 2021 | NL | Fig. 3D |

All modelling was performed in R (R Core Team, 2022). All data and code are archived at https://github.com/ahurford/pandemic-COVID-zero. Parameter estimates are summarized in Tables 2 and 3.

**Table 2:** Parameter estimates. All parameters are unitless.

| Description | Value | Details |
| --- | --- | --- |
| The number of close contacts infected per imported infection of the Original variant | $c_{OR}=0.149$ | Estimated from NLCHI data as: (the number of close contacts)/(the number of importations) on any day. Estimate is the average, weighted by the Original variant prevalence in Canada, when this prevalence was greater than 1%. |
| The number of close contacts infected per imported infection of the Alpha variant | $c_{\alpha}=0.114$ | Same estimation method and data as $c_{OR}$ |
| The number of close contacts infected per imported infection of the Delta variant | $c_{\delta}=0.266$ | Same estimation method and data as $c_{OR}$ |
| The number of close contacts infected per imported infection of the Omicron variant | $c_o=0.756$ | Same estimation method and data as $c_{OR}$ |
| Probability of no infection given exposure for unvaccinated individuals | $z_{0,k}=0$ | Assumed. Constraint is $0 \leq z_{0,k} \leq 1$. |
| Probability of no infection given exposure for individuals with 1 dose of vaccine (original, Alpha and Delta variant) | $z_{1,OR}=z_{1,\alpha}$<br>$=z_{1,\delta}$<br>$=0.49$ | Based on Pfizer vaccine efficacy against symptomatic infection (Bernal et al. 2021). Original variant same as Alpha variant (Khateeb et al. 2021). Constraint is $0 \leq z_{1,k} \leq 1$. |
| Probability of no infection given exposure for individuals with 1 dose of vaccine (Omicron variant) | $z_{1,o}=0$ | Assumed. Constraint is $0 \leq z_{1,k} \leq 1$. |
| Probability of no infection given exposure for individuals with 2 or 3 doses of vaccine (original and Alpha variant) | $z_{2,OR}=z_{2,\alpha}$<br>$=z_{3,OR}$<br>$=z_{3,\alpha}$<br>$=0.93$ | Based on Pfizer vaccine efficacy against symptomatic infection (Bernal et al. 2021). Original variant same as Alpha variant (Khateeb et al. 2021). Constraint is $0 \leq z_{2,k} \leq 1$. |
| Probability of no infection given exposure for individuals with 2 or 3 doses of vaccine (Delta variant) | $z_{2,\delta}=z_{3,\delta}$<br>$=0.88$ | Based on Pfizer vaccine efficacy against symptomatic infection (Bernal et al. 2021). Constraint is $0 \leq z_{2,k} \leq 1$. |
| Probability of no infection given exposure for individuals with 2 doses of vaccine (Omicron variant) | $z_{2,o}=0.09$ | Andrews et al. 2022. Based on Pfizer vaccine efficacy against symptomatic infection after 25 weeks. Constraint is $0 \leq z_{2,k} \leq 1$. |
| Probability of no infection given exposure for individuals with 3 doses of vaccine (Omicron variant) | $z_{3,o}=0.67$ | Andrews et al. 2022. Based on Pfizer vaccine efficacy against symptomatic infection. Constraint is $0 \leq z_{3,k} \leq 1$. |
| Transmission rate | $\beta = 0.287$ | Calibrated |
| Multiplicative change in transmission for Alpha variant relative to original variant | $b_{\alpha}=1.77$ | Model 1a estimate from Table 1 in Davies et al. 2021 |
| Multiplicative change in transmission for Delta variant relative to original variant | $b_{\delta}=1.97$ | Campbell et al. 2021 |
| Multiplicative change in transmission for Omicron variant relative to original variant | $b_o=2.97$ | Relative risk for unvaccinated primary cases (1.51) from Table 1 in Jalali et al. 2022 |
| Proportion of travellers that comply with self-isolation requirements | $\rho=0.7$ | Assumed. Constraint is $0 \leq \rho \leq 1$. |
| Probability the traveller is infectious after completing $x$ days of self-isolation | Figure S3E | See Supplementary Material for details |
| Probability a PCR test $a$ days after arrival is a false negative | Figure S3F | See Supplementary Material for details |
| Probability 5 Rapid Antigen Tests are all false negatives | $\sigma=0.1$ | Assumed. Constraint is $0 \leq \sigma \leq 1$. |

**Table 3:** Post-arrival travel restrictions in NL. The restrictions for travelers with 3 doses of vaccine are the same as for 2 doses of vaccine. For the calculations, 1 dose of vaccine and partially vaccinated were considered equivalent. When a vaccination status is not listed under a Special Measures Order (SMO), the new SMO does not change the measures that apply to that vaccination status. The functions *f*_1_(*s*) and *f*_2_(*s, τ*) are defined in the Supplementary Material.

| Reopening step | Dates, $t$ | New measures | Value |
| --- | --- | --- | --- |
| SMO (Travel Exemption Order), May 5 2020 | $t_1 = \text{May 4, 2020-June 30, 2021}$ | <u>All travellers</u> self-isolate for 14-days | $m_0(t_1) = f_1(14) = 0.153$<br>$m_1(t_1) = 0.153$<br>$m_2(t_1) = 0.153$ |
| SMO Reopening – Travel – Step 1, July 1, 2021 | $t_2 = \text{July 1, 2021-July 31, 2021}$ | <u>Partially vaccinated</u> travellers must have a negative PCR test result at entry<br><u>Fully vaccinated</u> travellers have no restrictions | $m_0(t_2) = 0.153$<br>$m_1(t_2) = f_2(0,0) = 0.384$<br>$m_2(t_2) = f_1(0) = 0.509$ |
| SMO Reopening – Travel – Step 2 - August 1, 2021 | $t_3 = \text{Aug 1, 2021-Sept 29, 2021}$ | Unvaccinated travellers complete a Polymerase Chain Reaction (PCR) test on day 7-9 of self-isolation, and can exit self-isolation if negative.<br><u>Partially vaccinated</u> travellers have no restrictions | $m_0(t_3) = 0.156$<br>$m_1(t_3) = f_2(8,8) = 0.509$<br>$m_2(t_3) = 0.509$ |
| SMO Reopening – Travel – Step 2 – UPDATED, Sept 30, 2021 | $t_4 = \text{Sept 30- Dec 20, 2021}$ | <u>Partially vaccinated</u> travellers complete a PCR test on day 7-9 of self-isolation, and can exit self-isolation if negative. | $m_0(t_4) = 0.156$<br>$m_1(t_4) = 0.156$<br>$m_2(t_4) = 0.509$ |
| SMO Reopening – Travel – Step 2 – December 21, 2021 Update | $t_5 = \text{Dec 21-24, 2021}$ | <u>All travellers</u> self-isolate for 5-days and complete a rapid antigen test each day | $m_0(t_5) = 0.1f_1(5) = 0.022$<br>$m_1(t_5) = 0.022$<br>$m_2(t_5) = 0.022$ |

### Modelling framework

Central to our approach where we develop a method to quantify the expected number of spillovers are two quantities: *n*_*j,k*_(*t*), the number of travel-related infections, and *p*_*j,k*_(*t*), the probability that a traveler or their close contacts infects a community member, where both quantities depend on the date, *t*. Travelers and their close contacts are indexed by their vaccination status, i.e., the number of vaccination doses completed, *j* which can be 0, 1, 2, or 3, and the infecting variant, *k*, referring to the Original (*OR*), Alpha (*α*), Delta (*δ*), or Omicron (*o*; BA.1 subvariant) variants. Post-arrival travel restrictions, NPIs and vaccine coverage in the local community, and variant transmissibility are all considered to calculate *p*_*j,k*_(*t*).

### Statistical model of imported cases to NL

To model the daily number of imported cases arriving to NL, we used a Poisson regression. Explanatory variables were time series of the mean new cases per 10,000 population over the last 14 days for (from east to west) Nova Scotia, Quebec, Ontario, Manitoba, Saskatchewan, Alberta and British Columbia. Provincial population sizes were based on Statistics Canada estimates for the first quarter of 2021. Fitted coefficients were constrained to be non-negative because we hypothesized that high infection prevalence in other provinces should have a positive relationship with the number of imported cases arriving in NL from that province.

### Model 1: Expected number spillovers

#### Characteristics of travel-related cases

To estimate the expected number of travelers or their close contacts that infected NL community members (referred to as ‘spillovers’), we first obtain the number of travelers and their close contacts with vaccination status *j* and infected with variant *k* as

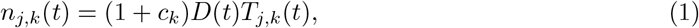

where *c*_*k*_ is the number of close contacts infected per imported infection of the variant, *k*, and *D*(*t*) is the number of imported infections reported on date *t*. We assume all infected travelers are identified and reported. Equation (1) assumes a similar frequency of vaccination statuses for travelers and their close contacts. Given a travel-related infection, *T*_*j,k*_(*t*) is the probability that the traveler has vaccination status *j* and is infected with variant *k*, where

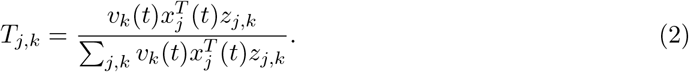

Here, 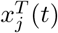 is the fraction of travelers with vaccination status *j* at time *t, v*_*k*_(*t*) is the frequency of the variant *k* at the origin sites of travelers, and *z*_*j,k*_ is the probability that a traveler with vaccination status *j* is infected with the variant *k*, where we assume no changes in *z*_*j,k*_ over time, i.e., as might occur due to waning of the vaccination.

As data on variant frequencies is not reliably available for jurisdictions within Canada, we parameterize *v*_*k*_(*t*) as the variant frequency in Canada. To parameterize *z*_*j,k*_, we equate reported vaccine efficacies against symptomatic infection with the probability of infection (see Table 2). Realistically, vaccines prevent less against infection and transmission than symptomatic infection, however, data for vaccine efficacies against infection and transmission are less available.

#### Post-arrival travel restrictions

Post-arrival travel restrictions may include self-isolation for a specified number of days after arrival, and Polymerase Chain Reaction (PCR) or Rapid Antigen Tests (RATs). In NL, different post-arrival travel restrictions were implemented through Special Measures Orders at different times during the SARS-CoV-2 public health emergency and depended on the vaccination status of the travelers (Table 3). We let *m*_*j*_(*t*) describe the efficacy of travel restrictions for a traveler with the vaccination status *j*, given the post-arrival travel restrictions on a given date *t*.

To estimate *m*_*j*_(*t*), we assumed that the efficacy of self-isolation for a given number of days could be calculated from the generation interval of SARS-CoV-2 (Ferretti et al., 2020), which was estimated for the Original variant. We assumed the generation interval was the same for all variants, although data suggests shorter generation times for the Delta variant (Hart et al., 2022). We felt this assumption was reasonable as our conclusions are likely more sensitive to other parameter estimates (as described in the Discussion). We estimated the probability of a false negative PCR test by considering Hellewell et al. (2021). The complete details of how we parameterized the effect of post-arrival travel restrictions are provided in the Supplementary Material. We had no information on compliance with self-isolation requirements, or when travelers are usually infected prior to arrival, and so we assumed 70% compliance with self-isolation, and that infected travelers were exposed between zero and ten days prior to arrival, with exposure times following a uniform distribution.

We assumed that the travel restrictions that applied to travelers also applied to their close contacts (i.e., household members). Our assumption is an over-simplification because in NL sometimes household members of travelers were subject to restrictions and other times they were not. If close contacts were infected from a traveler the timing of the close contact’s infectious period would be later than that of the traveler, and potentially after even a long period of self-isolation that began when the traveler arrived. This suggests that our assumption that the same restrictions apply to the traveler and their close contacts could under-estimate the spillover risk. However, in NL during the period of this study, if the traveler tested positive or if the close contacts developed symptoms, the close contacts were required to complete a PCR test. If the PCR test was positive, the close contacts were required to self-isolate, and in this respect, our assumptions regarding the probability that a close contact of a traveler infects a community member are an under-estimate.

#### NPIs and vaccination in the local community

We let the suspectibility of the local community to infection be determined by NPIs and vaccination (PIs). We let *ω*(*t*) be the stringency of NPIs in the local community on a given date *t*. We used the Bank of Canada COVID-19 stringency index estimated for NL. The Bank of Canada COVID-19 stringency index is calculated from 12 sub-indices which include policy related to school and workplace closures, restrictions on public and private gathering, travel restrictions, enforcement mechanisms, and public information campaigns (Cheung et al., 2021). We let *β* be a transmission rate parameter, and we let *b*_*k*_ be a multiplier reflecting the relative transmission rates for different variants.

The susceptibility of the local community to infection when considering vaccination is,

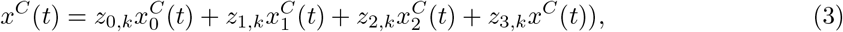

where the fraction of the community with different vaccination statuses is 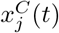 and *z*_*j,k*_ is the probability of infection given vaccination status *j* and the infecting variant *k* as previously defined. We assume four factors act independently to determine the probability that a traveller infects a local community member: (i) the efficacy of travel restrictions, *m*_*j*_(*t*); (ii) the stringency of NPIs, *ω*(*t*); (iii) the transmissibility of different variants; and (iv) the susceptibility of the local community after considering vaccination, *x*^*C*^(*t*). As such,

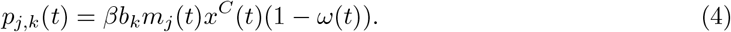

We assume that spillovers occur following a Binomial distribution with probability *p*_*j,k*_(*t*) and *n*_*j,k*_(*t*) trials. Then, on each date *t*, the expected number of community members infected by a traveler or their close contact is

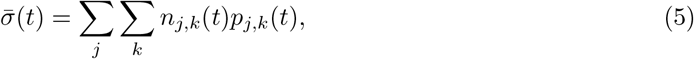

which is the expectation of a Binomial distribution summed across all vaccination and variant types.

This quantity, 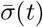, describes the daily expected number of community members infected by travelers and their close contacts (spillovers). Quantifying SARS-CoV-2 risks in regions that do not have SARS-CoV-2 community cases was an area of need during the first 18 months of the SARS-CoV-2 pandemic, and Equation 5 addresses this need.

### Model 2: Modelling outbreaks in regions implementing an elimination strategy

The second model for quantifying SARS-CoV-2 risk in regions that do not have community cases is to answer the question ‘if a community outbreak is established, how will the number of cases change over time, and how many cases will occur in the outbreak?’ To illustrate this modelling for a region that had few community cases of SARS-CoV-2, we consider Mount Pearl, NL.

Prior to December 15, 2021 in NL, the largest community outbreak of SARS-CoV-2 occurred due to the Alpha variant, with symptom onset dates from February 1 to 27, 2021, and with spread predominately in the Mount Pearl region. Mount Pearl is a suburb of St. John’s, and is part of the St. John’s metropolitan area which in 2016 had a population size of 205,955 (Statistics Canada, 2017). In response to the outbreak, on February 11, a Special Measures Order enacted the strictest level of NPIs (Alert level 5) in the St. John’s region. Contacts of cases were traced and tested, and many cases were associated with Mount Pearl Senior High School (Government of Newfoundland and Labrador, 2021b). No new cases associated with the outbreak were reported with symptom onset dates after February 28, 2021.

We calibrated a stochastic Susceptible-Infected-Recovered (SIR) model to data describing daily new reported cases and their symptom onset dates for cases belonging to the Mount Pearl outbreak (see Supplementary Material for details). This parameterized model is then the basis to explore the dynamics of hypothetical future outbreaks in Mount Pearl, NL.

For comparison, hypothetical scenarios retain the pattern of NPI implementation that occurred in the actual Mount Pearl outbreak, i.e., implementation of strict NPIs 10 days after the start of the outbreak, although it is possible to explore scenarios without this assumption. We consider future scenarios where vaccination coverage may have changed, and where a different variant may have caused the outbreak. For simplicity in interpreting the results of vaccination scenarios, we assume that all individuals in the community are either unvaccinated or have had two doses of vaccine.

## Results

Most of the SARS-CoV-2 cases reported in Atlantic Canada and Canada’s territories were travel-related from July 1, 2020 to May 31, 2021 (Figure 1). The period prior to July 1, 2020 was not considered because very few cases of any type were reported during this time. Notable differences that occur between these jurisdictions are that a much lower percentage of travel-related cases was reported each day in NB (mean = 36.7%, median = 13.4%), NT (mean = 12.8%, median = 0%) and YT (mean = 30.1%, median = 0%), as compared to NL (mean = 76.6%, median = 100%), NS (mean = 61.2%, median = 80%), and PE (mean = 91.3%, median = 100%) (Figure 1G). The values reported for NB are likely still much higher than the provinces west of NB, which had community spread and likely near 0% of reported cases were travel-related on most days.

**Figure 1:**
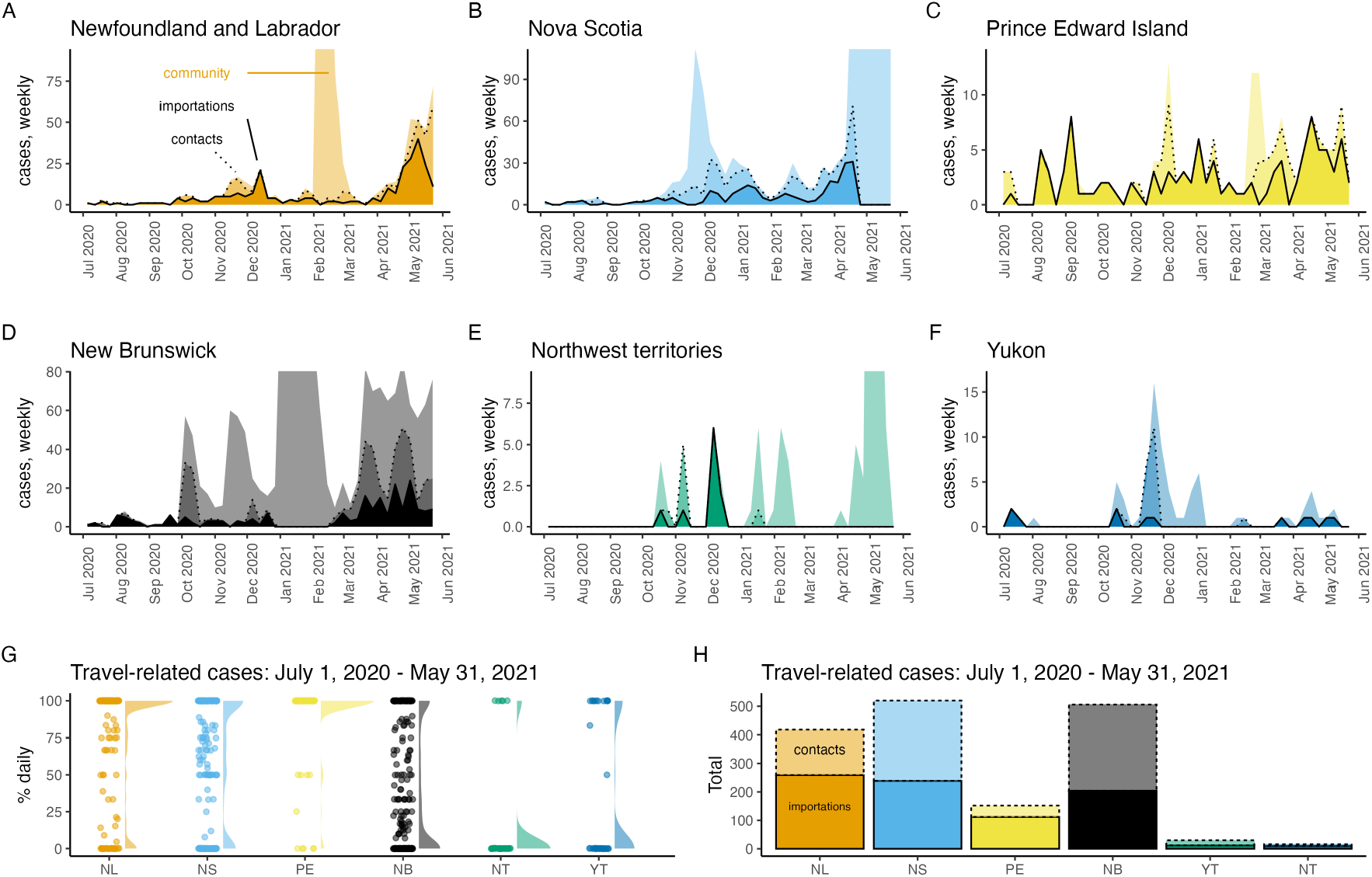
In Atlantic Canada and Canada’s territories most SARS-CoV-2 cases were importations and close contacts of these travelers from July 1, 2020 to May 31, 2021. Panels A-F show imported cases (dark shading, solid line), their close contacts (medium shading, dashed line), and community cases (light shading, no line) with the vertical axis limit as 20% more than the maximum number of reported weekly travel-related cases so that brief periods of large community outbreaks do not dominate the graphs. From July 1, 2020 – May 31, 2021, panels G-H show the percentage of reported daily cases that were travel-related (dots; also shown as a shaded density plot, G), and the total number of imported cases and their close contacts (H).

During the same period, the total number of travel-related cases also differed between jurisdictions with NL (importations = 259, close contacts of travellers = 159), NS (importations = 239, close contacts = 281), and NB (importations 204, close contacts = 302) having reported at least 2.75 times more travel-related cases than PE (importations = 112, close contacts = 40), and with YT (importations = 12, close contacts = 18) and NT (importations = 10, close contacts = 6) having reported very few travel-related cases at all (Figure 1H). Other Canadian provinces and Nunavut (NU) were not considered because travel-related case data was not reliably reported for these jurisdictions.

We found that the daily number of importations to NL was predicted as 1.12 times the mean number of new cases per 10,000 population in NS, where the mean is taken over the last 14 days (Figure 2). Estimated coefficients for the contribution of other provinces to the prediction of daily imported cases to NL were not different than zero, and the estimated intercept was zero. This statistical relationship is reasonable since a pre-pandemic survey reported 26% of travel into NL was from the Maritimes, second only to Ontario (Government of Newfoundland Labrador, 2018). The island of Newfoundland was the destination for 93% of travelers into NL (Government of Newfoundland Labrador, 2018), the ferry to Newfoundland departs from NS, and many flights to Newfoundland are routed with layovers in NS.

**Figure 2:**
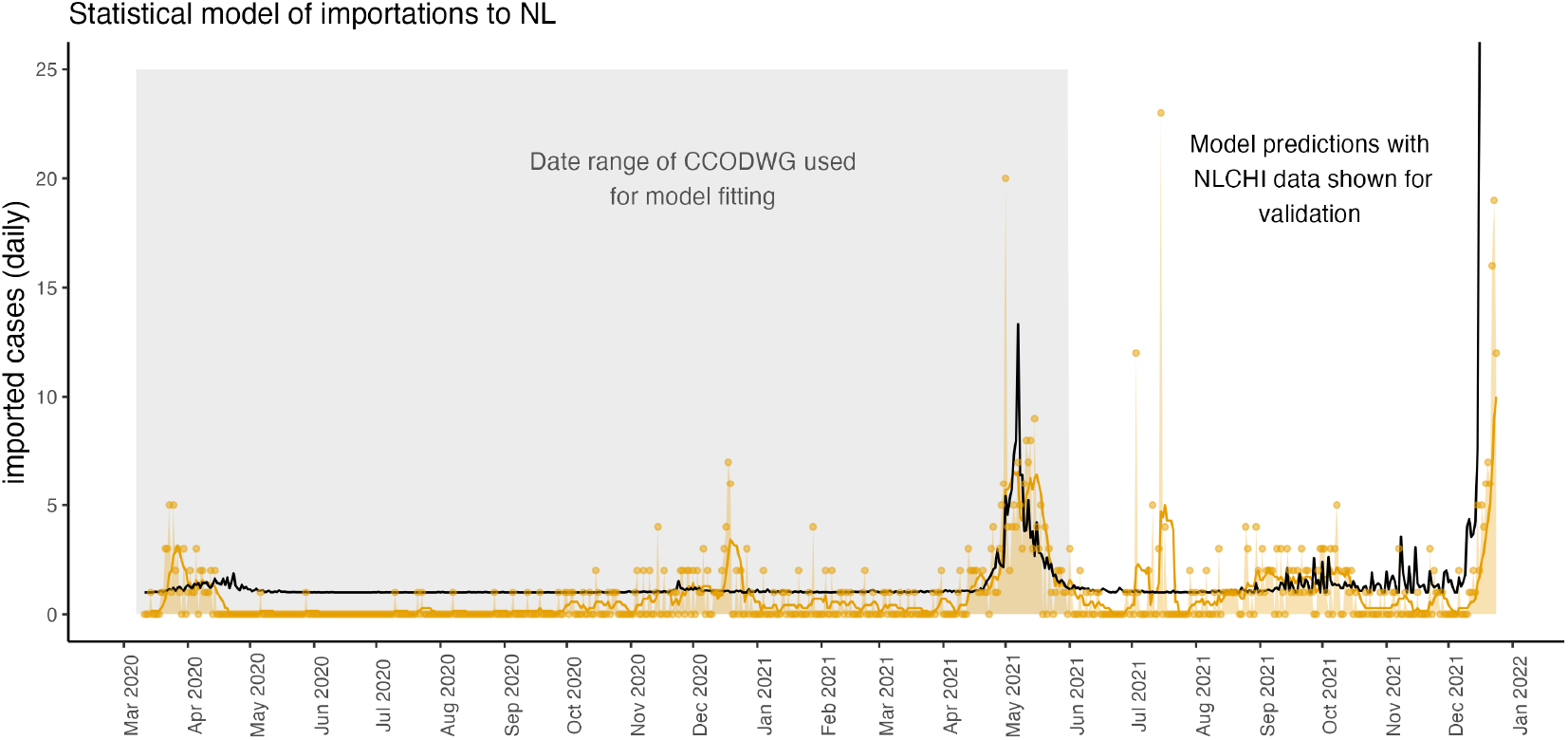
From March 15, 2020-December 24, 2021, the daily number of imported cases to NL is reliably predicted as 1.12 times the mean number of new cases per 10,000 population in NS, where the mean is taken over the last 14 days. This relationship was fit using the publicly available CCODWG data, where curation of these data ended on May 31, 2021 before the end of the study period. The model-predicted daily number of imported cases to NL (black line) extends beyond the time period of model fitting (grey shaded region) because data describing new cases in NS was available through to the end of the study period. To validate the predictions of the statistical model, we show the number of daily imported cases reported by NLCHI (yellow dots and yellow shading), and the 7-day rolling mean of daily imported cases (yellow line) where these data span the full study period.

The agreement of the model (Figure 2, black line) with the data (yellow line) is good since the model was only parameterized with data to May 31, 2021 (grey shaded region), but the model predictions still agree with the validation data from June 1 to December 24, 2021 and the model predicts the rise in importations that occurred in early December 2021. Few importations were reported and so chance events disrupt the agreement between the model predictions and the data. For example, in July 2021, a Portuguese fishing boat anchored in Conception Bay, NL and 31 crew members tested positive for SARS-CoV-2 (Smelie, 2021). This event may explain the 23 imported cases reported on July 15, 2021. The arrival of such boats with SARS-CoV-2 positive crew members is a chance event rather than a regularly occurring event that can be predicted by a model.

By December 24, 2021 (Figure 3E), the expected number of spillovers, infections spread from travellers and their close contacts to NL community members, was as high as it had ever been (as calculated by Equation 5). At this time, a community outbreak involving the Omicron variant was already occurring, with the first Omicron variant case in NL reported in St. John’s on December 15, 2021. The expected number of spillovers in mid-December was 44% higher than the previous highest value, and due to both the high number of imported cases (Figure 3A) and the reduced efficacy of two vaccine doses in protecting the NL community from infection with the Omicron variant (Figure 3C). In late July 2021, the expected number of spillovers was also high (Figure 3E). This was after NL relaxed entry requirements for Canadian travelers on July 1, 2021 (Figure 3B; Table 3), but before most Newfoundlanders and Labradorians were fully vaccinated (Figure S2C). The peak in the expected number of spillovers due to the Alpha variant (early May 2021; Figure 3E) was due to an increased number of importations occurring at that time (Figure 3A). The expected number of spillovers occurring due to the Delta variant was higher than that of the Alpha variant for two reasons: 1) after July 1, 2021 travel restrictions into NL for Canadians were relaxed (Figure 3B; Table 3), and 2) the Delta variant is more transmissible than the Alpha variant (Table 2).

**Figure 3:**
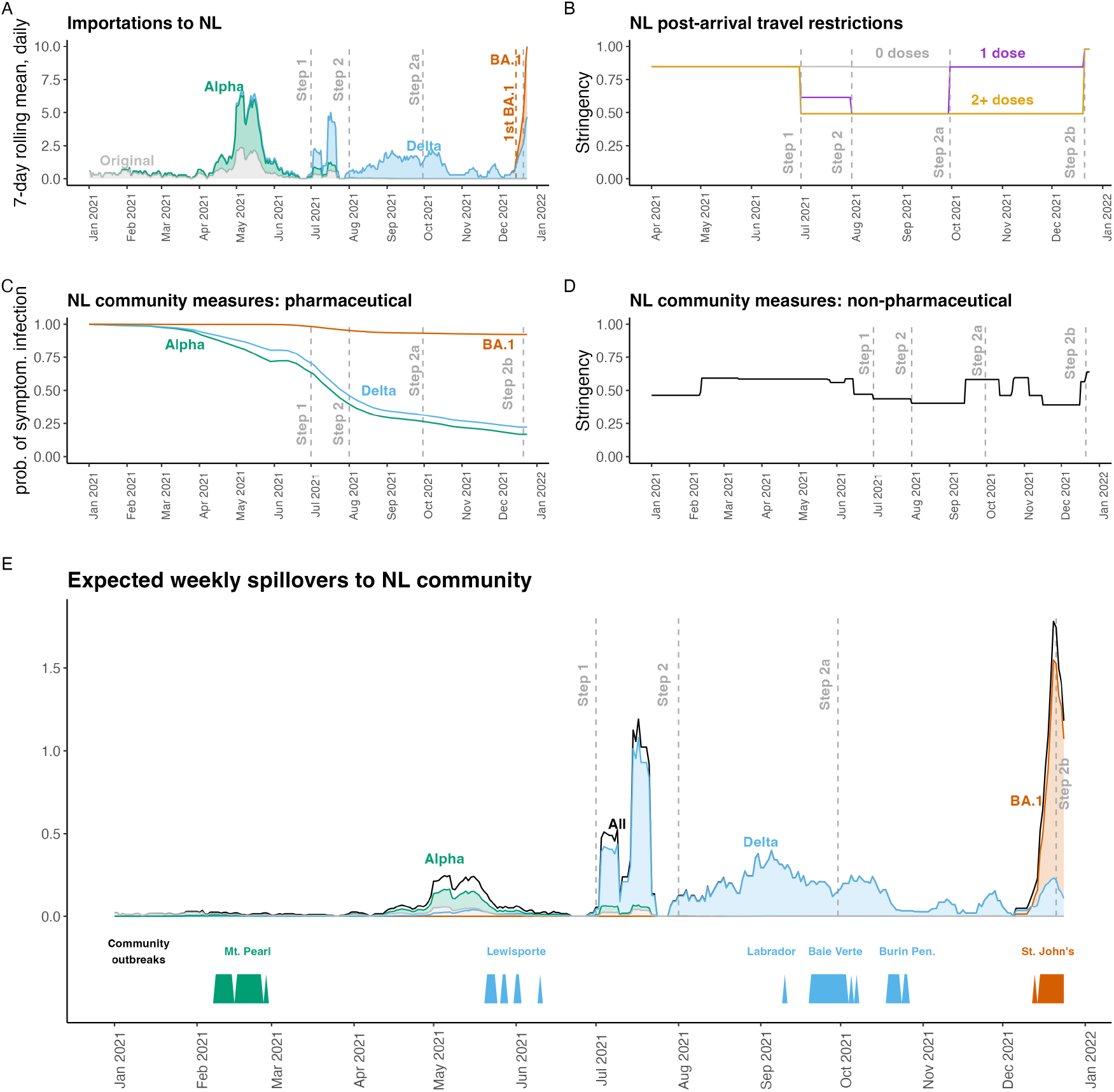
In mid-December 2021, the expected number of spillovers to NL community members was the highest it had ever been. High spillover risk in mid-December 2021 was due to the establishment of the Omicron BA.1 variant in Canada and high numbers of imported cases (A), and low vaccine efficacy for NL community members with two doses of vaccine exposed to the Omicron variant (C). Imported cases, *D*(*t*). (B) The stringency of post-arrival travel restrictions, 1 − *m*_*j*_ (*t*). (C) The probability of a symptomatic infection given exposure when considering vaccination of NL community members, *x*^*C*^(*t*). (D) The stringency of NPIs implemented in the NL community, *ω*(*t*). (E) The expected number spillovers, NL community members infected by travelers and their close contacts, 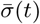 (black - equation 5; with variant-specific numbers shown with colours). The timing of actual community outbreaks with more than 5 cases are shown along the bottom bar. Grey dashed vertical lines show post-arrival travel restrictions due to different Special Measures Orders (see Table 3).

The stochastic SIR model (Figure 4A, green lines and shading) shows close agreement with the data from the Mount Pearl outbreak in February, 2021 (Figure 4A, green dots). When the Mount Pearl outbreak occurred few NL community members were vaccinated or had been infected, such that all the scenarios shown in Figure 4A assume a fully susceptible community. The Omicron variant (BA.1 subvariant) is much more transmissible than the other variants, and a hypothetical BA.1 variant outbreak in a fully susceptible Mount Pearl, NL community (Figure 4A, red line) cannot be completely shown given the y-axis limits that were set to emphasize the actual Mount Pearl Alpha variant outbreak. Figure 4A does not consider the arrival of imported cases. This is because the Mount Pearl data was strictly for cases known to belong to this outbreak.

**Figure 4:**
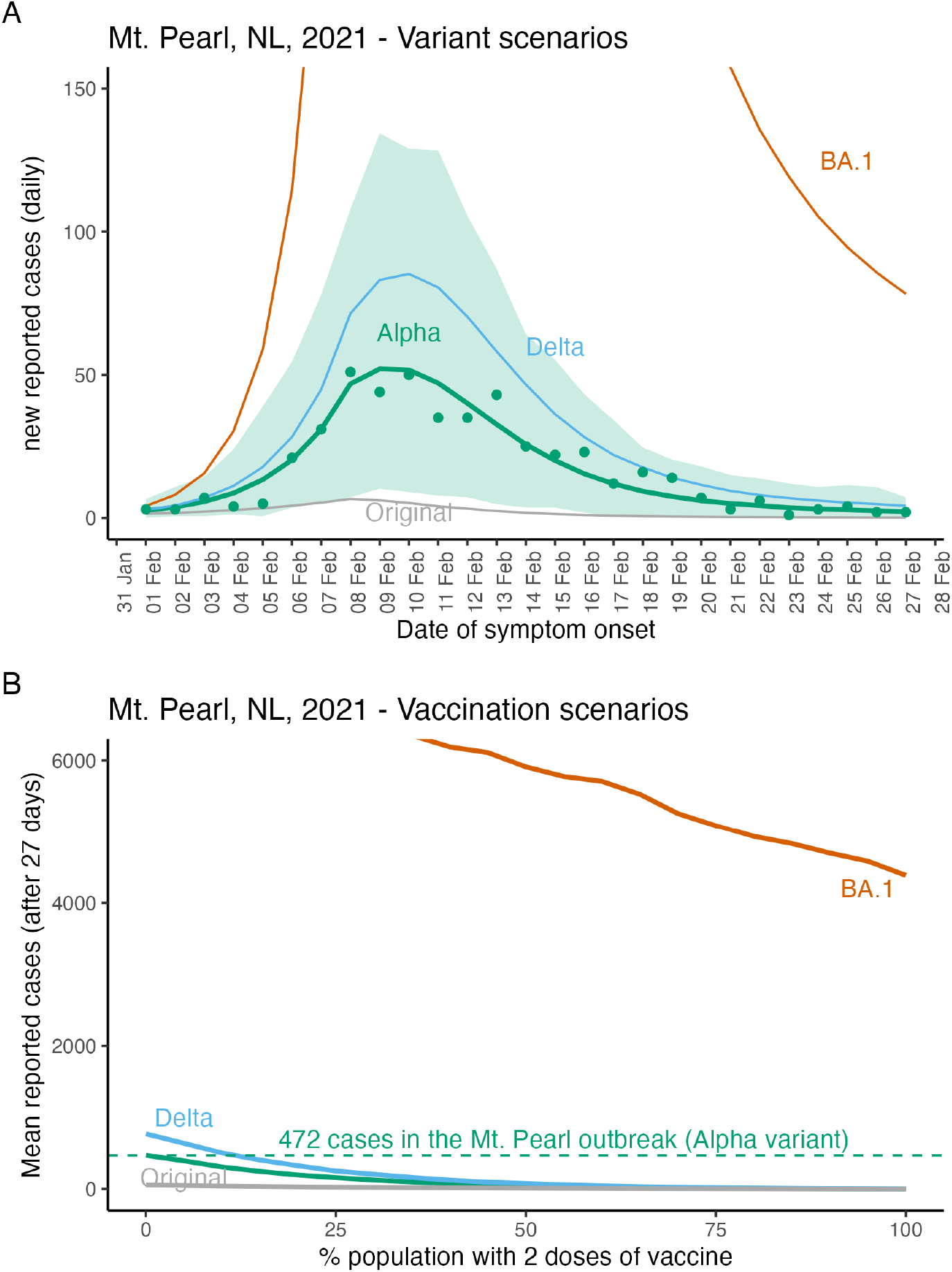
Epidemiological model fit and hypothetical future variant and vaccination scenarios for Mount Pearl, NL. The Mount Pearl outbreak was due to the Alpha variant and the y-axis limits of panel (A) were selected to show the Alpha variant (green line), and the Mount Pearl data (green dots) which meant that large values for the BA.1 variant are not shown. Lines show the mean and the shaded region shows the minimum and maximum values for 1000 simulations. The peak number of reported daily new cases for the BA.1 variant is 806 (not shown). In panel (B) vaccination scenarios assume community members are either unvaccinated or vaccinated with 2 doses. After 27 days of a BA.1 outbreak in a fully unvaccinated community, we estimate 7852 reported cases (not shown due to truncation). For more details describing parameter estimates see Table 2, and for model details see the Supplementary Material.

The number of cases reported in the Mount Pearl outbreak was 472 (Figure 4B, green dashed line). For the simulations, the mean total number of reported cases after 27 days (the duration of the Mount Pearl outbreak) when the community is fully susceptible are Original variant, 56, Alpha variant, 472, Delta, 773, and Omicron variant, 7852. We assumed that community members could be either unvaccinated or have two doses of vaccine. The effect of vaccination is to substantially reduce the number of reported cases in the outbreak after 27 days for all variants (Figure 4B).

## Discussion

In regions that have extended periods with few community cases of SARS-CoV-2, for example, regions that effectively implemented an elimination strategy, travel-related cases are a high percentage of reported cases (Arino et al., 2021; Godin et al., 2021), and modelling importations is particularly important (Zhang et al., 2022). Here, we extend such importation modelling to incorporate post-arrival travel restrictions, community vaccination coverage, and NPIs into the risk assessment frameworks for regions with few community SARS-CoV-2 infections.

Atlantic Canada and Canada’s territories experienced few SARS-CoV-2 cases prior to June 2021, however, there were some differences between these jurisdictions. NT and YT reported few travel-related cases, while NL, NS, and NB reported similar numbers of travel-related cases, but with NB reporting a much lower percentage of daily cases that were travel-related (Figure 1). Finally, while NL and NS had similar epidemiology until May 31, 2021, NL had enacted strict travel restrictions (Hurford et al., 2021), while NS enacted an extensive community testing program (Johnson-León et al., 2021). The YT implemented strict travel restrictions, but experienced an outbreak of the Gamma variant that overwhelmed hospital capacity (McPhee-Knowles et al., 2022).

We considered a statistical model describing the daily number of reported importations arriving in NL. During the pandemic response it was helpful to use this approach to forecast importations so that future risk could be assessed using Equation 5. That was not done in this manuscript because such an exercise would never be current, but this could be valuable to assess border measures, the threat of a new variant, or the impacts of waning immunity. We found that importations to NL could be predicted from the mean new cases per 10,000 people in NS over the last 14 days (Figure 2). These data were publicly available and regularly updated, but more generally better access to data describing travel volumes, travelers’ points of origin, reasons for travel, and granting of travel exemptions would aid real time importation modelling and risk quantification.

We applied our framework (Equation 5) to inform the potential for community outbreaks in NL. The estimated risk is somewhat consistent with the actual community outbreaks that occurred in NL (Figure 3). Generally, it seems difficult to predict when community outbreaks might occur in regions without community cases even given the vast amounts of data that were available during the SARS-CoV-2 pandemic.

Our analysis considers only known travel-related infections, such that estimates per infected traveler equate to per *known* infected traveler. In NL, for the pandemic until July 1, 2021, testing of arriving travelers was intensive (owing to few ports of entry, reduced travel volumes (Hurford et al., 2021), testing requirements for rotational workers (Department of Health and Community Services, NL, 2022), and requests for travelers potentially exposed during inbound flights to report for asymptomatic testing). This intensive testing, combined with few occurrences of community cases, suggests that a high proportion of imported cases were detected in NL during this time.

The main limitation of our analysis is parameter estimation and uncertainty. It is difficult to estimate the change in relative transmissibility due to a new variant because these data are estimated in different regions (or pooled across regions), and as the susceptible population changes owing to vaccination, infection, and waning of immunity during the time period that the estimation is made. We used 77% as the estimate of increased transmissibility of the Alpha variant relative to the Original variant (Table 2), however, the source of this estimate (Davies et al., 2021) gives a range values from 43% to 90% depending on the population and assumptions of the estimation procedure. Vaccine efficacies are estimated in specific populations, and application to other regions assumes no differences in population structure with regard to age and immunity, and does not estimate protection against infection and onward transmission, which is a critical parameter for epidemiological models. Finally, the impact of NPIs on transmission is difficult to assess, and the impact of new variant characteristics on the effectiveness of NPIs is unknown. In some instances data were not available to estimate parameters, for example, we assumed 70% compliance with self-isolation requirements, and the transmission rate parameter was calibrated (Table 2).

Our work was motivated by a need for regions that successfully implemented an elimination strategy during the first 18 months of the COVID-19 pandemic to quantify the risk of SARS-CoV-2 spread in their communities, and a need for guidelines to exit an elimination strategy when high vaccination coverage has been achieved. While guidelines for reopening have been developed by many jurisdictions, those using criteria expressed as the number of observed community cases (Anderson et al., 2021; Nali et al., 2021) are not helpful for regions that are reopening when there are few community cases.

Existing theory applicable to developing such guidelines is importation modelling (i.e., considering infection prevalence at travelers’ origins and travel volumes into a destination, e.g. Russell et al. 2021) and branching process modelling that calculates the probability of a major outbreak (i.e., Allen 2008). Extensions of classic branching process models consider the probability of an outbreak in age-structured populations with NPIs (Lovell-Read et al., 2022), and when importations occur (Ball et al., 2017). A related concept is the ‘event reproduction number’, a quantity that describes the number of secondary infections arising from one infected person attending an event (Tupper et al., 2020), since this quantity measures outbreak risk rather than simulating the entire outbreak. Some modelling studies have considered the efficacy of pre- and post-arrival travel restrictions (Steyn et al., 2021; Wells et al., 2021), but without linking to importation modelling as we have done. Future work to inform guidelines to exit an elimination strategy should further bridge these different research areas.

There is a need to communicate reasonable expectations to the public in regions where elimination has been implemented as relaxation of measures might have little or no impact on reported case numbers when infection prevalence is already high (Russell et al., 2021; Chen et al., 2020), but might bring risk in populations with zero or low SARS-CoV-2 prevalence (Russell et al., 2021; Chen et al., 2020; Arino et al., 2021). In regions that have implemented an elimination strategy, an increase in reported case numbers may occur even when measures are carefully and reasonably relaxed, and particularly if the prevalence of variants of concern is higher outside the jurisdiction than in (Wells et al., 2020; Grépin et al., 2021).

Prior to May 31, 2021, Atlantic Canada and Canada’s territories had experienced prolonged periods with few community SARS-CoV-2 cases. In this manuscript, we characterize differences within these jurisdictions, and distinguish between travel-related and community cases (Figure 1). We illustrate a type of epidemic modelling that is useful in these regions. This framework extends importation modelling such that border restrictions, variants, NPIs and vaccination in the local community are considered. Additionally, hypothetical future outbreaks are considered by simulating variant and vaccination scenarios. Our framework can be used to inform the risk associated with different candidate reopening plans when vaccination coverage is high in regions that have experienced prolonged periods with few SARS-CoV-2 cases, and help inform plans to exit an elimination strategy.

## Data Availability

All publicly available data used in this study are available at https://github.com/ahurford/pandemic-COVID-zero, but until the work completes peer-review please request these data and code by emailing

## Acknowledgements

AH, MM, FA, JW, BG and JA are supported by the Emerging Infectious Disease Modelling Consortium (the Canadian Network for Infectious Disease Modelling, Mathematics for Public Health, and the One Health Modelling Network for Emerging Infectious Diseases). AH was supported by a National Sciences and Engineering Research Council of Canada Discovery Grant (RGPIN 2014-05413) and funding from the Newfoundland and Labrador Department of Health and Community Services. AH acknowledges conversations with Proton Rahman, Sanjeev Sahara, and responding to modelling requests from the NL Department of the Health and Community Services that helped in the development of the modelling framework presented. NL COVID-19 data was provided by the NLCHI (Health Research Ethics Board reference number 2021.013).

## Supplementary Material

### Validation of the CCODWG data

The COVID-19 Canada Open Data Working Group (CCODWG; Berry et al. 2020 and Berry et al. 2021) is a group of volunteers who curate data from government and non-government sources. Data from the CCODWG line list are the basis of Figure 1. To validate these data, we compared the reported number of imported cases in the CCODWG data for Newfoundland and Labrador (NL) with the same data reported by the Newfoundland and Labrador Centre for Health Information (NLCHI).

Additionally, we compared the reported number of imported cases in the CCODWG data for New Brunswick (NB) with the same data reported from News Releases from the Government of New Brunswick (Government of New Brunswick, 2022). Each news release has a ‘new cases’ section that specifies the number of new cases and whether they are travel-related (which we define as an imported case in this manuscript), contacts of previously confirmed cases, or under investigation. For the most part, cases that are labelled ‘under investigation’ are not travel-related, since travel-related cases are quickly apparent. The news releases refer to the cases of the previous day and are recorded as occurring on the previous date. There is frequently a 1 day offset with the CCODWG data for this reason (corrected for in Figure S1).

**Figure S1:**
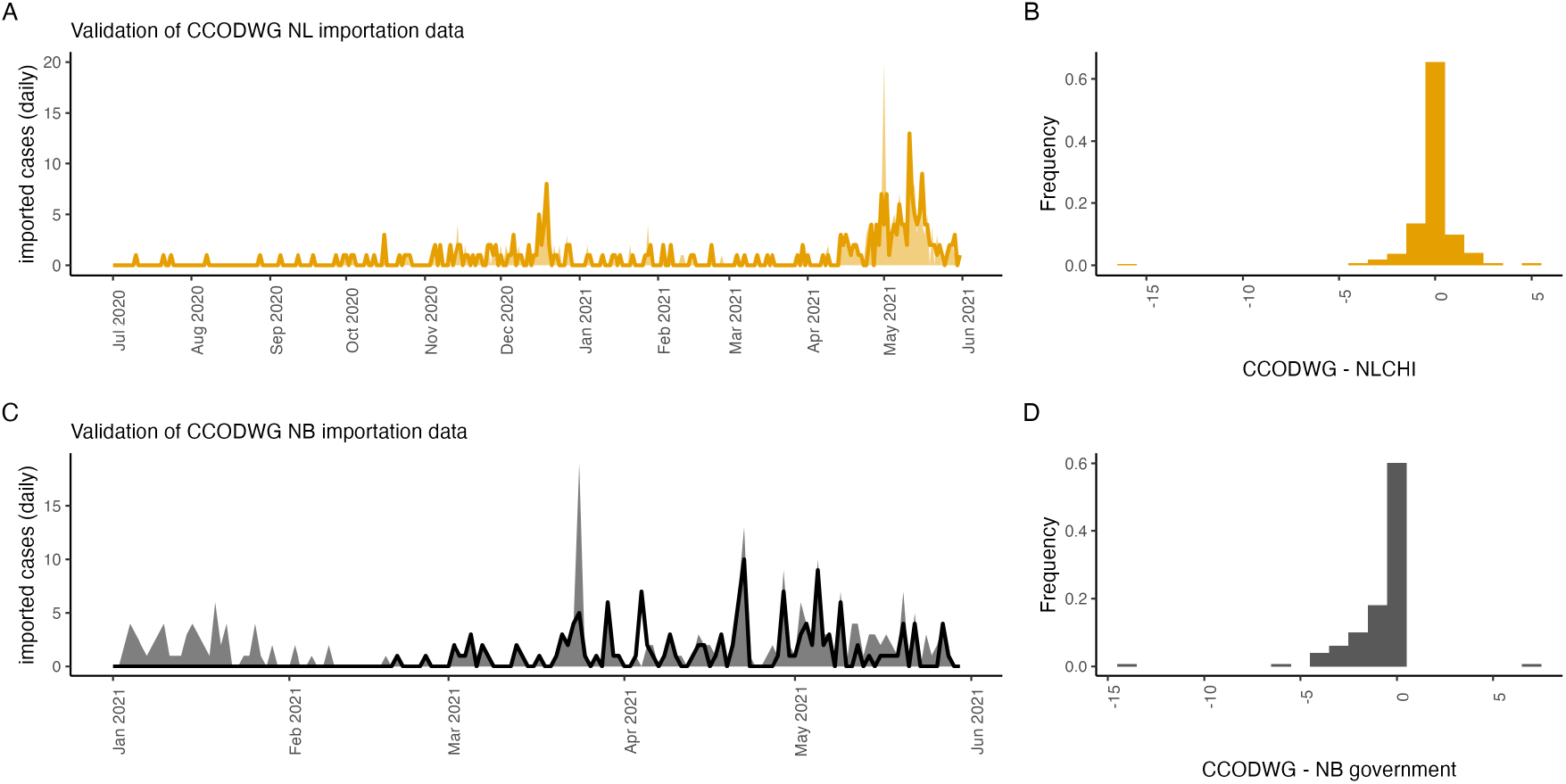
The daily number of imported cases reported in the CCODWG line list for NL and NB agrees with the number of imported cases reported by the NL and NB governments. (A) The CCOWDG daily reported importations for NL (orange line) compared to NLCHI data (orange shading). Frequency histogram of reported importations for NL: CCOWDG value - NLCHI values (July 1, 2020 – May 31, 2021). (C) The CCOWDG daily reported importations for NB (black line) compared to NB government reports (grey shading). D: Frequency histogram of reported importations for NB: CCOWDG value - NB government reported value (January 1, 2021 – May 31, 2021).

Figure S1 shows that the CCODWG data agrees with government data. Figure S1A and C show few differences between the CCOWDG data (lines) and the government data (shading). For both NL and NB over 60% of the daily reported number of imported cases are identical for both the CCOWDG data and the governments’ data (Figure S1B and D).

### Variant and vaccination data

The frequency of SARS-CoV-2 variants at the travelers’ points of origin *v*_*k*_(*t*) is assumed to be the variant frequency in Canada. As variant and vaccination data were reported weekly, we used linear interpolation to infer daily values. The variant data that we use is from the Public Health Agency of Canada (Figure S2A). These data are full genome sequences completed at the National Micro-biological Laboratory in Winnipeg, and from contributing provincial and territorial laboratories. Samples were contributed from provincial and territorial surveillance and sequencing priorities and volumes have changed over time. In August and September 2022, PHAC-reported variant frequencies were based on more than 3,000 genomes sequenced each week. The vaccination data that we use is from the Public Health Agency of Canada, and is shown in Figure S2B. The proportion of travelers with a given vaccination status, 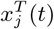, is assumed to be equal to the frequency of the different vaccination statuses in Canada (Figure S2B). We use the vaccination status of travelers to determine the applicable border restrictions (Equation 1). We use the vaccination status of Newfoundlanders and Labradorians to calculate the susceptibility of the NL population to spillover infections (Equation 3; Figure 3B).

**Figure S2:**
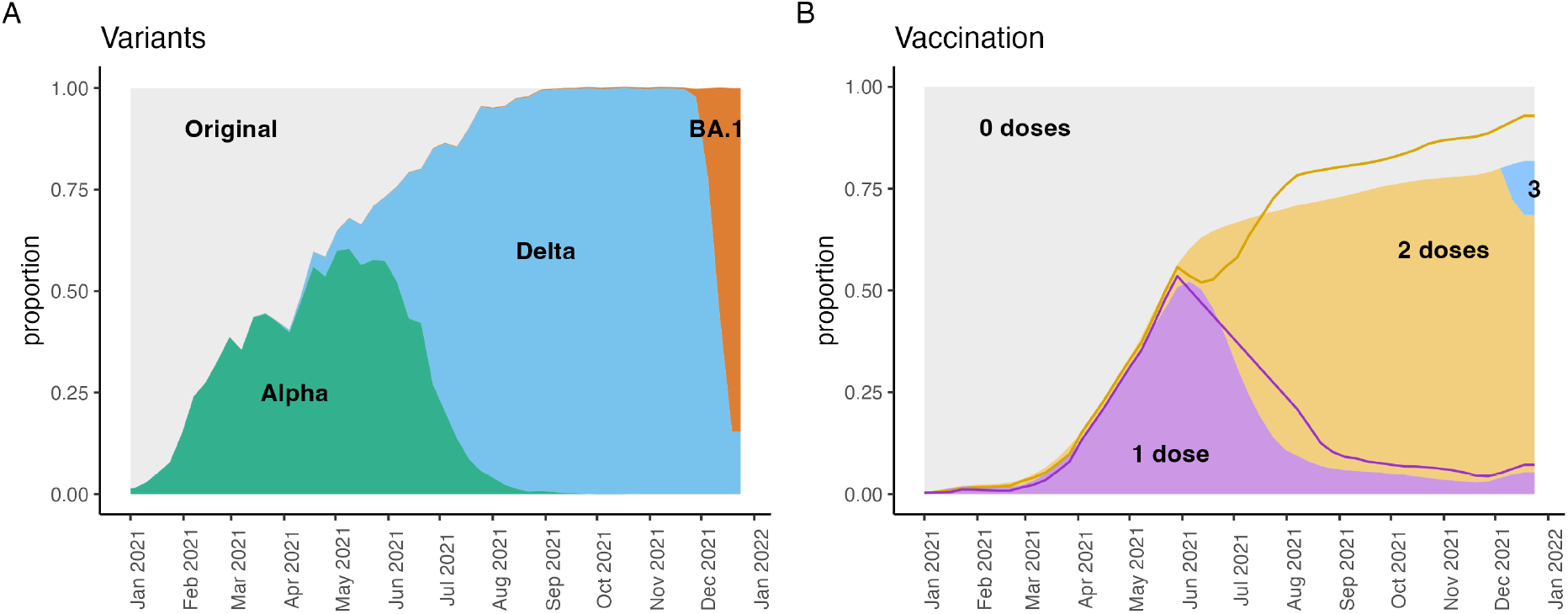
Variant proportions and vaccination coverage. (A) The proportion of genome sequences for each variant, *v*_*k*_(*t*). Samples were provided by Canadian provinces and territories. (B) The proportion of the population with different vaccination statuses for Canada (shaded; 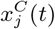) and NL (lines; 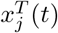). Vaccination statuses are unvaccinated, first doses, second doses, and third doses.

### Additional details for parameter estimation

To determine the impact of shorter duration self-isolation on the probability that a traveler infects a member of the NL community we ask, ‘how much potential infectivity remains if an individual leaves self-isolation after *n* days?’ This depends on when the traveler was infected (see Figure S3), and here we assume the number of days since infection of the arriving traveler is uniformly distributed between 0 to 10 days ago.

Infectivity as a function of days since exposure is based on Ferretti et al. (2020) who report that the generation interval for the Original variant follows a Weibull distribution with shape parameter equal to 2.83 and scale parameter equal to 5.67 (mean value of 5 days between infections). For self-isolation for *s* days, expected infectivity remaining is calculated as

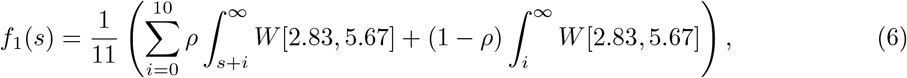

where W[2.83,5.67] is the Weibull distribution for the generation interval and *ρ* is the probability of compliance with self-isolation. Equation 6 is illustrated in Figure S3. Note that even if no self-isolation is required the fraction of infectivity remaining is only 0.51 owing to the assumption of potential exposure prior to entry.

If a PCR test is required as part of the travel restrictions, then we consider the probability of a true positive (the test sensitivity) as based on Figure 3B in Hellewell et al. (2021):

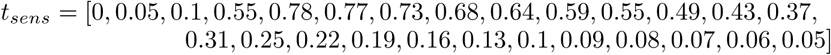

where these values correspond to 0 to 25 days since exposure (see Figure S3). The probability of a false negative is 1− *t*_*sens*_ and is multiplied by the infectivity remaining after any self-isolation to estimate the expected infectivity of a traveler in the community:

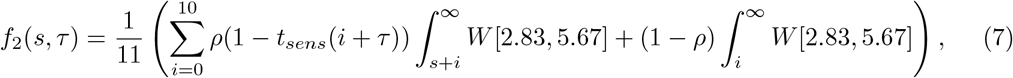

where *τ* is number of days after arrival when the PCR test is performed, and it is assumed that only travelers that were complying with self-isolation would also self-isolate given a positive test result.

**Figure S3:**
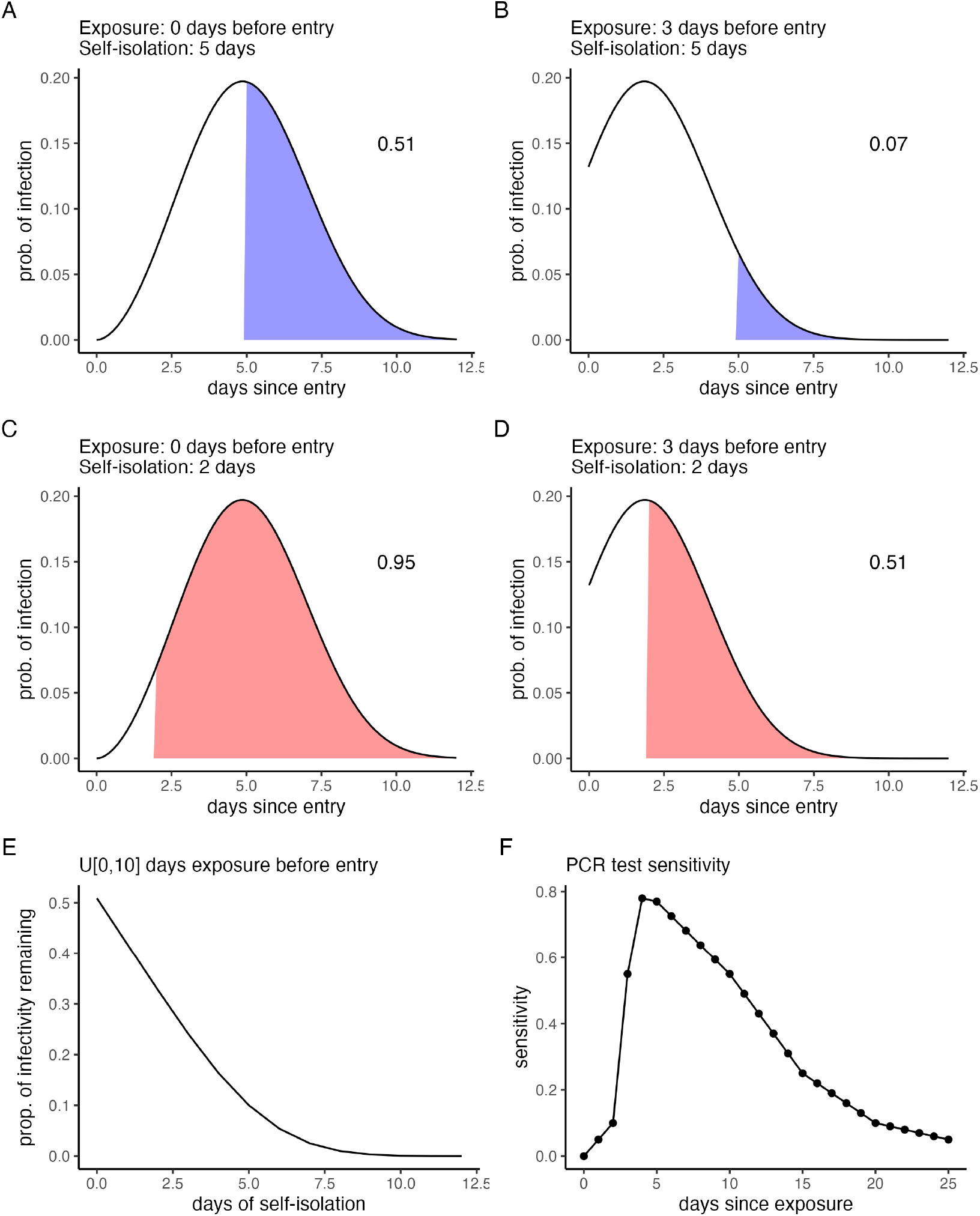
Graphical illustration of the parameter estimation method for post-arrival travel restrictions. Panels A-D show 4 combinations of days since exposure at entry and durations of self-isolation (blue: 5 days and red: 2 days), where the shaded area (also provided as a numerical value in the panel) is the infectivity remaining after the traveler exits self-isolation. (E) The expected proportion of infectivity remaining when assuming the days since exposure at entry is uniformly distributed between 0 and 10 days. (F) Polymerase Chain Reaction test specifity, *t*_*sens*_.

### Stochastic SIR model

A stochastic Susceptible-Infected-Recovered (SIR) model was calibrated to fit the Mount Pearl, NL outbreak data. This was achieved by changing the value of the transmission rate, *β*_0_, until the total number of cases predicted by the model for the Alpha variant when there was no vaccination in the community was within 1 case of the observed total number of cases for the Mount Pearl outbreak: 472.

The stochastic SIR model is a Markov chain for the time interval (*t, t* + ∆*t*] the transition probabilities are

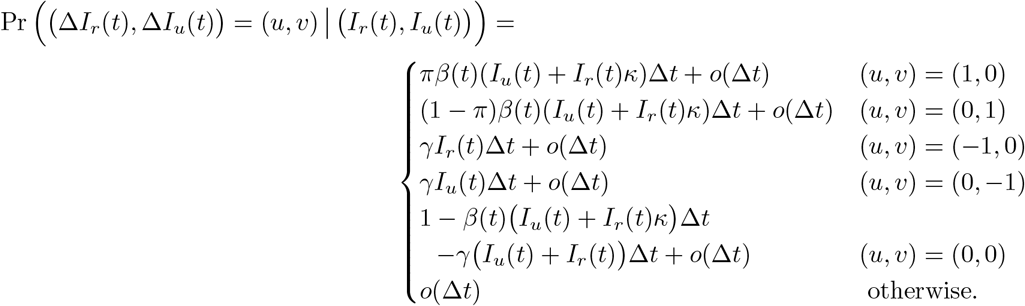

where *I*_*r*_(*t*) and *I*_*u*_(*t*) are the number of reported and unreported cases actively infected at time *t*, respectively. The probability that an infection is reported is *π* (assumed to be 0.6). The fraction of infected individuals that self-isolate is *κ* (assumed to be 0.5). The recovery rate from infection is *γ* (assumed to be 0.1 days). The transmission rate is *β*(*t*) and is assumed to decrease from *β*_0_ to *a*_1_*β*_0_ after some time *t*\* (assumed to be 10 days after the outbreak begins). We assume the change in *β*(*t*) occurs gradually, at an exponentially decreasing rate, with coefficient *a*_2_, such that,

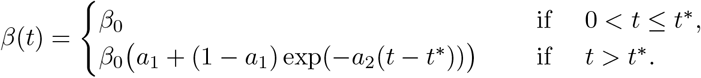

Our model can be expressed as a multivariate counting process **N**(*t*) with exponential inter-arrival times and intensity,

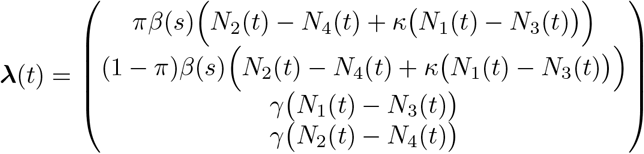

where *N*_*j*_(*t*), *j* ∈ {1, 2, 3, 4} are the counts up to time *t* of reported infections, unreported infections, and individuals recovered from the reported and unreported groups, respectively.

We do not consider the number of susceptible individuals as a dynamic variable since for the application of this model to regions that have few community cases of SARS-CoV-2 it is likely that the outbreak infects only a very small fraction of the population. Therefore, the number of susceptible individuals remains relatively constant. We assume that vaccination coverage does not change during the outbreak because the outbreak is relatively short.

Parameter values for the calibrated fit are *β*_0_ = 0.887, *a*_1_ = 0.03, and *a*_2_ = 0.325.

The effect of different variants was considered by multiplying *β*_0_ by the appropriate values given in Table 2. The effect of vaccination was considered by multiplying *β*_0_ by *x*_0_ + *x*_2_*z*_2,*k*_ where *x*_0_ is the fraction of the population that is unvaccinated, *x*_2_ is the fraction of the population that is fully vaccinated (assumed to be 1 − *x*_0_), and *z*_2,*k*_ is the efficacy of two doses of vaccine against symptomatic infection (assumed to be equivalent to infection; see Table 2). Only *β*_0_ was changed for the different hypothetical outbreaks.

### NL reopening plan: Together.Again

**Table S1:** NL’s reopening plan: Together.Again. Table 3 describes the actual course of reopening in NL, which deviated from the reopening plan.

|  | Travel | Non-pharmaceutical interventions | Vaccination targets |
| --- | --- | --- | --- |
| **Transition**<br>**(June 15 – July 1)** | • 14-day isolation is required for all individuals entering NL.<br>• Only NL residents, asymptomatic workers, individuals subject to the Exemption order, and individuals allowed entry due to extenuating circumstances approved by the Chief Medical Officer of Health are permitted entry to NL.<br>• Individuals must be available for contact by public health following arrival. | • Non-medical masking required in indoor public spaces.<br>• Physical distancing.<br>• If potentially symptomatic, isolate and test.<br>• Receive both doses of COVID-19 vaccine.<br>• Outdoor gatherings limited to 150 people.<br>• Outdoor ceremonial events permitted with social distancing.<br>• Personal outdoor gatherings of up to 30 people.<br>• Personal indoor gatherings limited to 20 people |  |
| **Step 1**<br>**(July 1)** | • Non-essential travel permitted within Canada.<br>• Testing, self-isolation not required for fully vaccinated Canadians.<br>• Partially vaccinated Canadians require a negative test result, or must self-isolate until one is obtained.<br>• 14-day self-isolation required for unvaccinated Canadians. | • Non-medical masking required in indoor public spaces.<br>• Physical distancing.<br>• If potentially symptomatic, isolate and test.<br>• Receive both doses of COVID-19 vaccine.<br>• Outdoor gatherings limited to 250 people.<br>• Indoor gatherings limited to 200 people or 75% capacity.<br>• Personal outdoor gatherings of up to 50 people.<br>• Personal indoor gatherings limited to 20 people. | • D1: 75% |
| **Step 2**<br>**(August 15)** | • Testing, self-isolation not required for fully and partially vaccinated Canadians.<br>• Unvaccinated Canadians will be required to test on day 7, 8 or 9, and must self-isolate until a negative test result. | • Non-medical masking required in indoor public spaces.<br>• Physical distancing.<br>• If potentially symptomatic, isolate and test.<br>• Receive both doses of COVID-19 vaccine.<br>• Outdoor gatherings limited to 500 people.<br>• Indoor gatherings limited to 350 people.<br>• Personal gatherings restricted to number of people that can fit in the space while abiding by distancing regulations. | • D1: 80%<br>• D2: 50% |
| **Step 3**<br>**(Sept. 15)** | • Testing, self-isolation not required for fully and partially vaccinated Canadians.<br>• Unvaccinated Canadians must self-isolate until a negative test result. | • Masking guidelines will be decided based on current evidence.<br>• Physical distancing.<br>• If potentially symptomatic, isolate and test.<br>• Receive both doses of COVID-19 vaccine.<br>• No capacity restrictions on outdoor gatherings.<br>• Increased indoor gathering capacities.<br>• Personal gatherings restricted to number of people that can fit in the space while abiding by distancing regulations. | • D2: 80% |

## References

Michael G Baker, Nick Wilson, and Tony Blakely. Elimination could be the optimal response strategy for covid-19 and other emerging pandemic diseases. BMJ, 371, 2020a. doi: 10.1136/bmj.m4907.

Anita E Heywood and C Raina Macintyre. Elimination of COVID-19: what would it look like and is it possible? The Lancet. Infectious Diseases, 20(9):1005, 2020. doi: 10.1016/s1473-3099(20)30633-2.

Simona Bignami. The burden of COVID-19 in Canada. Canadian Studies in Population, 48(2): 123–129, 2021. doi: 10.1007/s42650-021-00056-w.

Damien Contandriopoulos. The year public health lost its soul: a critical view of the COVID-19 response. Canadian Journal of Public Health, 112(6):970–972, 2021. doi: 10.17269/s41997-021-00583-8.

Department of Health and Community Services, NL. Report to the house of assembly on the covid-19 public health emergency. September 14, 2022. URL https://www.assembly.nl.ca/business/electronicdocuments/ReporttoHOACOVID-19PublicHealthEmergency2022.pdf/.

Michael G. Baker, Nick Wilson, and Andrew Anglemyer. Successful elimination of Covid-19 transmission in New Zealand. The New England Journal of Medicine, 383:e56, August 2020b. ISSN 1533-4406. doi: 10.1056/NEJMc2025203. URL https://www.semanticscholar.org/paper/c279c1aff07a1adb4af655b8d0bf47c98cacc1f2.

Nguyen Hai Nam, Pham Nguyen Quy, Truong-Minh Pham, and Joel Branch. No new community COVID-19 infection in four consecutive weeks: what lesson can be learned from Vietnam. Journal of infection in developing countries, 14(10):1125–1127, October 2020. ISSN 1972-2680. doi: 10.3855/jidc.13080.

Lara B. Aknin, Bernardo Andretti, Rafael Goldszmidt, John F. Helliwell, Anna Petherick, Jan-Emmanuel De Neve, Elizabeth W. Dunn, Daisy Fancourt, Elkhonon Goldberg, Sarah P. Jones, Ozge Karadag, Elie Karam, Richard Layard, Shekhar Saxena, Emily Thornton, Ashley Whillans, and Jamil Zaki. Policy stringency and mental health during the COVID-19 pandemic: a longitudinal analysis of data from 15 countries. The Lancet. Public Health, 7:e417.–e426, May 2022. ISSN 2468-2667. doi: 10.1016/S2468-2667(22)00060-3.

Michael König and Adalbert Winkler. The impact of government responses to the covid-19 pandemic on gdp growth: Does strategy matter? PLOS ONE, 16:e0259362, 2021.

Government of Newfoundland and Labrador. News releases. public advisory: Update on covid-19 in newfoundland and labrador, december 31. Department of Health and Community Services, 2021a. URL https://www.gov.nl.ca/releases/2021/health/1231n06/.

Statistics Canada. Population estimates, Quarterly. Government of Canada, 2021. URL https://www150.statcan.gc.ca/t1/tbl1/en/tv.action?pid=1710000901.

Public Health Agency of Canada. COVID-19 epidemiology update. 2022. URL https://health-infobase.canada.ca/covid-19/.

Arisina Ma and Jane Parry. When Hong Kong’s “dynamic zero” covid-19 strategy met omicron, low vaccination rates sent deaths soaring. BMJ (Clinical research ed.), 377:o980. April 2022. ISSN 1756-1833. URL 10.1136/bmj.o980.

Andrew Silver. Covid-19: What went wrong after initial success in Laos? British Medical Journal, 377, 2022. URL doi:10.1136/bmj.o994.

Maria M. Martignoni and Amy Hurford. It’s not realistic to eliminate COVID-19 in Newfoundland and Labrador. Here’s why. Canadian Broadcasting Corportation, Newfoundland and Labrador, 2022. URL https://www.cbc.ca/news/canada/newfoundland-labrador/covid-19-here-to-stay-1.6384033.

Timothy W Russell, Joseph T Wu, Sam Clifford, W John Edmunds, Adam J Kucharski, Mark Jit, et al. Effect of internationally imported cases on internal spread of COVID-19: a mathematical modelling study. The Lancet Public Health, 6(1):e12–e20, 2021. doi: 10.1016/s2468-2667(20)30263-2.

Julien Arino, Nicolas Bajeux, Stephanie Portet, and James Watmough. Quarantine and the risk of COVID-19 importation. Epidemiology & Infection, 148:e298, 2020. doi: 10.1017/S0950268820002988. URL https://dx.doi.org/10.1017/S0950268820002988. See https://github.com/julien-arino/covid-19-importation-risk for code.

Tiange Chen, Siwan Huang, Guanqiao Li, Yuan Zhang, Ye Li, Jinyi Zhu, Xuanling Shi, Xiang Li, Guotong Xie, and Linqi Zhang. An integrated framework for modelling quantitative effects of entry restrictions and travel quarantine on importation risk of COVID-19. Journal of biomedical informatics, 118:103800, June 2021. ISSN 1532-0480. doi: 10.1016/j.jbi.2021.103800.

Borame L. Dickens, Joel R. Koo, Jue Tao Lim, Haoyang Sun, Hannah E. Clapham, Annelies Wilder-Smith, and Alex R. Cook. Strategies at points of entry to reduce importation risk of COVID-19 cases and reopen travel. Journal of Travel Medicine, 27, December 2020. ISSN 1708-8305. doi: 10.1093/jtm/taaa141.

David J Price, Freya M Shearer, and Michael T Meehan. Early analysis of the australian covid-19 epidemic. eLife, 9:e58785, 2020. URL https://doi.org/10.7554/eLife.58785.

Sarah P. Otto, Troy Day, Julien Arino, Caroline Colijn, Jonathan Dushoff, Michael Li, Samir Mechai, Gary Van Domselaar, Jianhong Wu, David J. D. Earn, and Nicholas H. Ogden. The origins and potential future of SARS-CoV-2 variants of concern in the evolving COVID-19 pandemic. Current biology : CB, 31:R918–R929, July 2021. ISSN 1879-0445. doi: 10.1016/j.cub.2021.06.049.

J.L. Juul, K. Græsbøll, and L.E. Christiansen. Fixed-time descriptive statistics underestimate extremes of epidemic curve ensembles. Nat. Phys., 17:5–8, 2021. URL https://doi.org/10.1038/s41567-020-01121-y.

Committee for the Coordination of Statistical Activities. How COVID-19 is changing the world: a statistical perspective. Technical report, United Nations, March 2021. Volume III. New York, 2021. https://unstats.un.org/unsd/ccsa/documents/.

Kamalini Lokuge, Emily Banks, Stephanie Davis, Leslee Roberts, Tatum Street, Declan O’Donovan, Grazia Caleo, and Kathryn Glass. Exit strategies: optimising feasible surveillance for detection, elimination, and ongoing prevention of COVID-19 community transmission. BMC medicine, 19 (1):1–14, 2021. doi: 10.1186/s12916-021-01934-5.

Open Society Common Purpose Taskforce. A roadmap to reopening. Technical report, University of Sydney, 2021.

Isha Berry, Jean-Paul R. Soucy, Ashleigh Tuite, and David Fisman. Open access epidemiologic data and an interactive dashboard to monitor the COVID-19 outbreak in Canada. Canadian Medical Association Journal, 192:E420, April 2020. ISSN 1488-2329. doi: 10.1503/cmaj.75262.

I. Berry, M. O’Neill, and S.L. et al Sturrock. A sub-national real-time epidemiological and vaccination database for the COVID-19 pandemic in Canada. Sci Data, 2021.

Calista Cheung, Jerome Lyons, Bethany Madsen, Sarah Miller, and Saarah Sheikh. The Bank of Canada COVID-19 stringency index: measuring policy response across provinces. 2021. URL https://www.bankofcanada.ca/2021/02/staff-analytical-note-2021-1/.

Nick Andrews, Julia Stowe, Freja Kirsebom, Samuel Toffa, Tim Rickeard, Eileen Gallagher, Charlotte Gower, Meaghan Kall, Natalie Groves, Anne-Marie O’Connell, David Simons, Paula B. Blomquist, Asad Zaidi, Sophie Nash, Nurin Iwani Binti Abdul Aziz, Simon Thelwall, Gavin Dabrera, Richard Myers, Gayatri Amirthalingam, Saheer Gharbia, Jeffrey C. Barrett, Richard Elson, Shamez N. Ladhani, Neil Ferguson, Maria Zambon, Colin N.J. Campbell, Kevin Brown, Susan Hopkins, Meera Chand, Mary Ramsay, and Jamie Lopez Bernal. Covid-19 vaccine effectiveness against the Omicron (B.1.1.529) variant. New England Journal of Medicine, 386(16): 1532–1546, 2022. doi: 10.1056/NEJMoa2119451.

Jasmin Khateeb, Yuchong Li, and Haibo Zhang. Emerging SARS-CoV-2 variants of concern and potential intervention approaches. Critical Care, 25(1):1–8, 2021. doi: 10.1186/s13054-021-03662-x.

Jamie Lopez Bernal, Nick Andrews, Charlotte Gower, Eileen Gallagher, Ruth Simmons, Simon Thelwall, Elise Tessier, Natalie Groves, Gavin Dabrera, Richard Myers, et al. Effectiveness of COVID-19 vaccines against the B. 1.617. 2 variant. medRxiv, 2021. doi: 10.1101/2021.05.22.21257658.

Nicholas G. Davies, Sam Abbott, Rosanna C. Barnard, Christopher I. Jarvis, Adam Kucharski, James D. Munday, and Carl B. Pearson. Estimated transmissibility and impact of SARS-CoV-2 lineage B.1.1.7 in England. Science, 372, 2021.

N. Jalai, Hilde K. Brustad, Arnoldo Frigessi, Emily A. MacDonald, Hinta Meijerink, Siri L. Feruglio, and Karin M. Nygard. Increased household transmission and immune escape of the SARS-CoV-2 Omicron compared to Delta variants. Nature Communications, 13, 2022.

R Core Team. R: A language and environment for statistical computing. R Foundation for Statistical Computing, Vienna, Austria, 2022. URL https://www.R-project.org/.

Luca Ferretti, Chris Wymant, Michelle Kendall, Lele Zhao, Anel Nurtay, Lucie Abeler-Dörner, Michael Parker, David Bonsall, and Christophe Fraser. Quantifying SARS-CoV-2 transmission suggests epidemic control with digital contact tracing. Science, 368(6491), 2020. doi: 10.1101/2020.03.08.20032946.

William S. Hart, Elizabeth Miller, Nick J Andrews, Pauline Waight, Philip K. Maini, Sebastian Funk, and Robin N. Thompson. Generation time of the alpha and delta SARS-CoV-2 variants: an epidemiological analysis. Lancet Infectious Diseases, 22:603–610, 2022.

Hellewell et al. Estimating the effectiveness of routine asymptomatic PCR testing at different frequencies for the detection of SARS-CoV-2 infections. BMC Medicine, 19, 2021. doi: 10.1186/s12916-021-01982-x.

Statistics Canada. Census profile, 2016 census. 2017. URL https://www12.statcan.gc.ca/census-recensement/2016/dp-pd/prof/index.cfm?Lang=E.

Government of Newfoundland and Labrador. Public advisory: 100 new cases of covid-19 in newfoundland and labrador. February, 2021, 2021b. URL https://www.gov.nl.ca/releases/2021/health/0211n05/.

Government of Newfoundland Labrador. 2016 exit survery - research highlights. 2018. URL https://www.gov.nl.ca/tcar/files/2016_Exit_Survey_Highlights_Report_FINAL_REVISED_June_2018.pdf.

Sarah Smelie. Third ship anchored off Newfoundland coast with COVID-19 cases among crew. The Globe and Mail, page July 19, 2021.

Julien Arino, Pierre-Yves Bö;elle, Evan M Milliken, and Stephanie Portet. Risk of COVID-19 variant importation – How useful are travel control measures? Infectious Disease Modelling, 6:875–897, 2021. doi: 10.1016/j.idm.2021.06.006. URL https://doi.org/10.1016/j.idm.2021.06.006.

Arnaud Godin, Yiqing Xia, David L. Buckeridge, Sharmistha Mishra, Dirk Douwes-Schultz, Yannan Shen, Maxime Lavigne, Mélanie Drolet, Alexandra M. Schmidt, Marc Brisson, and Mathieu Maheu-Giroux. The role of case importation in explaining differences in early SARS-CoV-2 transmission dynamics in Canada - a mathematical modeling study of surveillance data. International Journal of Infectious Diseases, 102:254–259, January 2021. ISSN 1878-3511. doi: 10.1016/j.ijid.2020.10.046.

Xianghong Zhang, Yunna Song, Sanyi Tang, Haifeng Xue, Wanchun Chen, Lingling Qin, Shoushi Jia, Ying Shen, Shusen Zhao, and Huaiping Zhu. Models to assess imported cases on the rebound of COVID-19 and design a long-term border control strategy in Heilongjiang Province, China. Mathematical Biosciences and Engineering, 19:1–33, January 2022. ISSN 1551-0018. doi: 10.3934/mbe.2022001.

Amy Hurford, Proton Rahman, and J. Concepción Loredo-Osti. Modelling the impact of travel restrictions on COVID-19 cases in Newfoundland and Labrador. Royal Society Open Science, 8: 202266, June 2021. ISSN 2054-5703. doi: 10.1098/rsos.202266.

Maureen Johnson-León, Arthur L Caplan, Louise Kenny, Iain Buchan, Leah Fesi, Phoebe Olhava, Desmond Nsobila Alugnoa, Mara G Aspinall, Emily Costanza, Brianna Desharnais, et al. Executive summary: It’s wrong not to test: The case for universal, frequent rapid COVID-19 testing. EClinicalMedicine, 33, 2021.

S McPhee-Knowles, B Hoffman, and L. Kanary. The Yukon’s experience with COVID-19: Travel restrictions, variants and spread among the unvaccinated. Can Commun Dis Rep, 48:17–21, 2022. URL https://doi.org/10.14745/ccdr.v48i01a03.

Sean C Anderson, Nicola Mulberry, Andrew M Edwards, Jessica E Stockdale, Sarafa A Iyaniwura, Rebeca C Falcao, Michael C Otterstatter, Naveed Z Janjua, Daniel Coombs, and Caroline Colijn. How much leeway is there to relax COVID-19 control measures? Epidemics, 35:100453, 2021. doi: 10.1016/j.epidem.2021.100453.

Luiz Henrique da Silva Nali, Felipe Scassi Salvador, Graciela dos Santos Soares Bonani, Heitor Franco de Andrade, Expedito José de Albuquerque Luna, and Dennis Minoru Fujita. Reopening borders: protocols for resuming travel during the COVID-19 pandemic, 2021.

Linda J. S. Allen. An introduction to stochastic epidemic models. In F Brauer, P van den Driessche, and J Wu, editors, Mathematical Epidemiology, chapter 3, pages 81–130. Springer, 2008.

Francesca A. Lovell-Read, Silvia Shen, and Robin N. Thompson. Estimating local outbreak risks and the effects of non-pharmaceutical interventions in age-structured populations: SARS-CoV-2 as a case study. Journal of Theoretical Biology, 535:110983, 2022.

Frank Ball, Tom Britton, and Pieter Trapman. An epidemic in a dynamic population with importation of infectives. The Annals of Applied Probability, 27:242–274, 2017.

Paul Tupper, Himani Boury, Madi Yerlanov, and Caroline Colijn. Event-specific interventions to minimize COVID-19 transmission. Proceedings of the National Academy of Sciences, 17:32038–32045, 2020.

Nicholas Steyn, Michael J. Plank, Alex James, Rachelle N. Binny, Shaun C. Hendy, and Audrey Lustig. Managing the risk of a COVID-19 outbreak from border arrivals. Journal of the Royal Society Interface, 18:20210063, 2021.

Chad R. Wells, Jeffrey P. Townsend, Abhishek Pandey, Seyed M. Moghadas, Gary Krieger, Burton Singer, Robert H. McDonald, Meagan C. Fitzpatrick, and Alison P. Galvani. Optimal COVID-19 quarantine and testing strategies. Nature Communications, 12:356, 2021.

Shi Chen, Qin Li, Song Gao, Yuhao Kang, and Xun Shi. State-specific projection of COVID-19 infection in the United States and evaluation of three major control measures. Scientific Reports, 10(1):1–9, 2020. doi: 10.1101/2020.04.03.20052720.

Chad R. Wells, Pratha Sah, Seyed M. Moghadas, Abhishek Pandey, Affan Shoukat, Yaning Wang, Zheng Wang, Lauren A. Meyers, Burton H. Singer, and Alison P. Galvani. Impact of international travel and border control measures on the global spread of the novel 2019 coronavirus outbreak. Proceedings of the National Academy of Sciences of the United States of America, 117:7504–7509, March 2020. ISSN 1091-6490. doi: 10.1073/pnas.2002616117.

Karen Ann Grépin, Tsi-Lok Ho, Zhihan Liu, Summer Marion, Julianne Piper, Catherine Z Worsnop, and Kelley Lee. Evidence of the effectiveness of travel-related measures during the early phase of the COVID-19 pandemic: a rapid systematic review. BMJ Global Health, 6(3):e004537, 2021. doi: 10.1136/bmjgh-2020-004537.

Government of New Brunswick. News Releases. 2022, 2022. URL https://www2.gnb.ca/content/gnb/en/corporate/promo/covid-19/news.html.

